# LabWAS: novel findings and study design recommendations from a meta-analysis of clinical labs in two independent biobanks

**DOI:** 10.1101/2020.04.08.19011478

**Authors:** Jeffery A. Goldstein, Joshua S. Weinstock, Lisa A. Bastarache, Daniel B. Larach, Lars G. Fritsche, Ellen M. Schmidt, Chad M. Brummett, Sachin Kheterpal, Goncalo R. Abecasis, Joshua C. Denny, Matthew Zawistowski

**Affiliations:** Department of Pathology, Northwestern University Feinberg School of Medicine, Chicago IL 60611, USA; Department of Biostatistics and Center for Statistical Genetics, University of Michigan School of Public Health, Ann Arbor, MI 48109, USA; Vanderbilt Genetics Institute, Vanderbilt University Medical Center, Nashville, TN 37232, USA; Departments of Medicine and Biomedical Informatics, Vanderbilt University Medical Center, Nashville, TN 37232, USA; Department of Anesthesiology, Keck School of Medicine, University of Southern California, Los Angeles, CA 90033, USA; Open Targets, Wellcome Sanger Institute, Hinxton, Cambridge, CB10 1SD, United Kingdom; Department of Anesthesiology, Michigan Medicine, University of Michigan, Ann Arbor, MI 48109 USA; All of Us Research Program, NIH (work completed while at Vanderbilt University Medical Center)

## Abstract

Phenotypes extracted from Electronic Health Records (EHRs) are increasingly prevalent in genetic studies. EHRs contain hundreds of distinct clinical laboratory test results, providing a trove of health data beyond diagnoses. Such lab data is complex and lacks a ubiquitous coding scheme, making it more challenging than diagnosis data. Here we describe the first large-scale cross-health system genome-wide association study (GWAS) of EHR-based quantitative lab measurements. We meta-analyzed 70 labs matched between the BioVU cohort from the Vanderbilt University Health System and the Michigan Genomics Initiative (MGI) cohort from Michigan Medicine. We show high replication of known association for these labs, validating EHR-based measurements as high-quality phenotypes for genetic analysis. Notably, our analysis provides the first replication for 700 previous GWAS associations across 46 different labs. We discovered 31 novel associations at genome-wide significance for 22 distinct labs, including the first reported associations for two labs. We replicated 22 of these novel associations in an independent tranche of BioVU samples. The summary statistics for all association tests are available through an interactive webtool to benefit other researchers. Finally, we performed mirrored analyses in BioVU and MGI to assess competing analytic practices for lab data. We find that using the mean of all available lab measurements provides a robust summary value, but alternate summarizations can improve power in certain labs. This study provides a proof-of-principle for cross health system GWAS and is a framework for future studies of quantitative traits in EHRs.

## Introduction

Laboratory testing is a key component of modern medicine. Lab measurements provide a glimpse into the functioning of the human body, allowing clinicians to diagnose and monitor disease. In most health systems, lab measurements are routinely captured in patient Electronic Health Records (EHRs) alongside disease diagnoses, free text notes and medical procedures to provide a detailed, longitudinal health history ^1^. EHRs present exciting research potential by providing broad phenotyping on large cohorts with minimal cost^2,3^.

Several large-scale genetic studies have already used EHRs as the source of phenotypes ^4–6^, most commonly based on International Classification of Diseases (ICD) codes mapped to dichotomous phenotypes^7^. Although disease is often thought of in all-or-nothing binary state, many diseases exist on a continuum with the ultimate clinical diagnosis occurring once a relevant quantitative lab measurement exceeds a pre-determined threshold. For example, hypercholesteremia, diabetes mellitus and chronic kidney disease are each diagnosed almost entirely on lab tests for low density lipoprotein (LDL), glycated hemoglobin (or glucose) and creatinine, respectively. The lab values are therefore a more sensitive measure of underlying health than diagnosis and may provide a more powerful analysis. As an example, hypercholesterolemia and coronary artery disease risk locus *PSCK9* was initially discovered based on quantitative LDL measurement rather than clinical diagnosis ^8,9^. In contrast to binary disease phenotypes, there are fewer examples of genetic analyses of EHR-derived quantitative lab values^10–12^.

This rich data source of quantitative lab measurements in large cohorts comes with unique concerns: Quantitative traits collected specifically for research purposes typically use a controlled experimental design to ensure consistency among samples. In contrast, lab values contained in EHRs are a historical record of medical care. As such, patients may have hundreds of lab measurements for some traits and none for others, depending on their specific health problems and utilization of the health system. The measurements can be collected in times of sickness or good health leading to substantial variation in values for the same lab. Lab measurements can be artificially modified by prescription medicine, such as statin use for LDL cholesterol. Moreover, recruitment mechanisms and demographics of a biobank can dramatically shape the overall health of the cohort, which in turn dictate lab measurements available for analysis. The impact of using such “real world” measurements for genetic association studies is unclear. Questions remain over the effect and robustness of analytic choices made when analyzing EHR-based labs including how best to summarize complicated, longitudinal lab measurements and whether diseases highly correlated with lab measurements should be considered. Prior studies are not consistent in addressing these concerns. For example, a genome-wide analysis of EHR-derived quantitative traits in Biobank Japan enrolled patients with at least 1 of 47 diagnoses and controlled for all 47 diagnoses while testing each lab ^13^. On the opposite end of the spectrum, an analysis of labs within the Geisinger EHR did not control for underlying disease states ^14^. The variety of methods to summarize lab values and models to test for genetic association indicates that the question of how best to handle these data remains unsettled.

In this paper we explore strategies for analyzing quantitative lab values extracted from EHRs and describe the first large-scale meta-analysis of EHR-derived lab traits across independent health systems. We used lab measurements and genetic data from two academic health systems: the BioVU cohort from Vanderbilt University^15^ and the Michigan Genomics Initiative (MGI) from Michigan Medicine^16^. Meta-analysis offers a mechanism to increase sample size and power for detecting genetic risk variants but comes with distinct challenges for EHR lab traits including matching labs between health systems and determining specific analysis plan for the complicated lab data. The cohorts differ dramatically in their recruitment mechanisms, patient composition and recording format for lab measurements: MGI was predominantly recruited through inpatient surgical encounters at Michigan Medicine whereas BioVU recruitment required outpatient appointments at Vanderbilt University Medical Center. As a result, MGI is enriched for diseases treated surgically such as extreme obesity and solid tumors. This heterogeneity reflects the reality of EHR-based phenotyping, and strategies must be developed for future collaborative work on the growing number of EHR-linked biobanks.

Our initial challenge was identifying which labs to meta-analyze between the health systems. Accurately matching labs is complicated by the fact that no standardized coding scheme exists for lab measurements. Dichotomous disease traits are easily matched between health systems using the ubiquitous ICD coding system for disease diagnoses^17^. Although the Logical Observation Identifiers Names and Codes (LOINC) system offers the promise of interoperability for lab traits, it is cumbersome and maps poorly onto other ontologies^18^. For example, there are 21 distinct codes for blood glucose which might not be used consistently between institutions. Health systems may instead adopt their own idiosyncratic internal terminology for electronic recording of labs. Based on a methodical manual review of EHR text descriptions and lab values, we identified 70 lab traits between BioVU and MGI that could be matched with high confidence. We extracted previously identified variants for these lab traits from the GWAS catalog to serve as true positive variants for assessing subsequent analyses. Our meta-analysis replicated nearly 75% true positive GWAS catalog variants, validating both the accuracy of lab matches across health systems and the overall quality of the EHR lab data. Further, we discovered 31 novel lab-associated variants across 22 labs, including the first reported associations for the saliva and pancreatic enzyme amylase and bicarbonate CO2, a gaseous waste product from metabolism carried in the blood. We immediately replicated 22 (71%) of these novel associations using an independent second set of BioVU samples.

The meta-analysis of the complicated lab data required several strategic choices regarding data preparation and statistical analysis. Using a series of mirrored analyses performed in MGI and BioVU, we explored the consistency of various analytic choices between the biobank cohorts. Specifically, we considered the statistic used to summarize individual-level lab values for the GWAS (mean, median, maximum or first available value) and the inclusion of disease covariates in the GWAS regression. We determined that although there is no single best strategy for analyzing labs, using the mean lab value not controlling for potentially relevant diseases proved reasonably robust strategy between MGI and BioVU across lab traits.

Our study represents a proof-of-principle for accurate matching and meta-analysis of quantitative lab measurements extracted from diverse EHRs. Just as the first wave of GWAS studies was followed by a wave of meta-analyses, we predict that meta-analysis of EHR-derived data is imminent. Our results indicate that, despite the heterogeneous demographics of health systems and recording of clinical data, meta-analysis between EHRs stands to be a powerful strategy for genetic discovery.

## Methods

### Datasets

We analyzed data from two university hospital biobanks that link electronic health records with genetic data: BioVU from Vanderbilt University and the Michigan Genomics Initiative (MGI) from Michigan Medicine. We restricted our analysis to unrelated patients of European ancestry because of insufficient patient sample sizes from non-European populations.

The BioVU cohort has been described previously^15^. Briefly, DNA was extracted from surplus blood samples and genotyping data was linked to de-idenified EHR data. For this study, we used a cohort of 20,515 individuals genotyped on the Multi-Ethnic Genotyping Array (MEGA) from Illumina and estimated to be of European ancestry by admixture^19^. We included 843,242 SNPs that passed standard marker QC filters and had a minor allele frequency >1%. We retrieved all available lab measurements in this cohort that occurred when the subject was at least 18 years of age.

The MGI cohort has also been described previously^16^. Briefly, MGI samples were recruited primarily through surgical encounters at Michigan Medicine and provided consent for linking of their EHRs and genetic data for research purposes. MGI samples were genotyped on customized Illumina HumanCoreExome v12.1 bead arrays. European samples were identified using Principal Component Analysis. We used a data freeze consisting of 40K unrelated European individuals for this analysis. MGI samples were imputed to the Haplotype Reference Consortium using the Michigan Imputation Server^20^, providing ∼14 million SNPs with a minimac imputation quality R2>0.3 and an allele frequency greater than 1e-6. We analyzed the set of ∼800K overlapping SNPs between the MGI imputed genotypes and the BioVU MEGA array for this study.

### Harmonization of Labs Between Health Systems and the GWAS Catalog

We extracted all available clinical lab measurements and metadata from the electronic health records of MGI samples and BioVU samples. The MGI lab data consisted of >31 million distinct lab measurements for 5098 unique lab names. We collapsed distinct labs when obvious duplications were present (e.g., “Eosinophils” and “EOSINOPHILS”). Available metadata differed slightly between the health systems but included brief text descriptions, unit of measurements, and range for normal values. We excluded individual lab measurements labelled as “External” and taken outside the health system. In cases where multiple tests examined the same analyte, e.g. blood glucose, we removed point of care (POC) tests which are more susceptible to technical artifacts and tend to be deployed in intensive care or emergency settings where acute disease or treatment effects supervene determinants of the underlying baseline ^21,22^.

We matched lab tests recorded at Vanderbilt and Michigan health systems based on manual curation of the metadata including recorded lab names, clinical descriptions, measurement units, range of measurements, and patient count. We selected a set of 70 labs matched with high confidence between the health systems and having at least 1,000 individuals with the lab measured in each cohort for further analysis.

### Disease phenotypes

In order to study the effect of underlying health conditions we extracted ICD9 and ICD10 diagnosis codes from the EHR of the BioVU and MGI cohorts. We searched for diagnosis for 42 diseases with the potential to alter a clinical lab measurement (Supplementary Table). We started with the disease list used in the BioBank Japan lab analysis^10^ and removed diseases which do not occur in our population (e.g. febrile seizures of infancy) and those expected to have minimal effect on labs (e.g. cataracts). We supplemented their list with chronic diseases expected to have a large impact on labs due to their prevalence (e.g. hypertension). We created an indicator variable for each disease (1 if the sample had at least one qualifying ICD code for the specific disease and a 0 otherwise) to include as covariates in GWAS regression analyses.

### Statistical Analysis

#### Intra-cohort Genome-wide Association Studies

We first performed GWAS analysis of each lab trait separately in the MGI and BioVU cohorts. To determine the impact of study design choices, we performed multiple GWAS for each lab. We varied the statistic used to summarize the longitudinal lab measurements available for each sample (mean, median, first available measurement and maximum available measurement), and whether indicators for disease diagnosis were included as covariates in the GWAS regression (yes/no).

For each GWAS, the distribution of lab summary statistics was inverse normalized separately within the MGI and BioVU cohorts prior to regression analysis. In a separate analysis of the BioVU cohort, we determined that inverse normalization of lab values performed better than applying no transformation, or a log or square root transformation for controlling GWAS type I error (data not shown). Genome-wide association tests were performed on the inverse normalized traits using additive linear regression models. We included age, sex and four principal components as covariates in each regression. The model controlling for disease status included an additional 42 covariates for the binary disease phenotypes. The regression analyses were performed in the BioVU cohort using PLINK^23^ and in the MGI cohort using *epacts 3*.*3*.*0* ^24^.

We treated the GWAS of mean trait value with no disease covariates as the default analysis and compared each alternate analysis to this default. We quantified the impact of each analysis strategy relative to the default analysis by computing the log fold change in p-value between the alternative and default analysis for each analyzed SNP. That is, for each SNP we compute the quantity

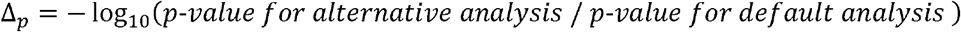

for the MGI analysis and the BioVU analysis separately. A positive value for Δ_*p*_ indicates a SNP that increases in significance (smaller p-value) when the alternate summary statistic is used and vice versa. We used scatterplots to display the simultaneous change in p-value for both cohorts, performing LD-pruning on non-catalog SNPs to simplify the pictures (See Figure 4). Since most SNPs are not associated with the lab trait of interest, alternative summarizations simply result in independent noise between the two cohorts, resulting in a diamond shaped pattern centered at the origin.

We implemented a heuristic to formally distinguish the SNPs with largest changes in p-value between the alternative and default analysis methods from those with movement due simply to random noise. The heuristic generates a bounding polygon around the diamond cluster of points. The polygon is generated by fitting a diamond to the set of Δ_*p*_ values, the joint distribution of the log fold changes in the two cohorts, using simulated annealing to estimate the shape of the diamond such that 99.9% of all SNPs are included within the boundaries. We defined SNPs outside the boundaries of the polygon as those with largest simultaneous changes in p-values in both cohorts. Catalog SNPS located outside the bounding polygon were classified as having either a concordant increased effect if p-value significance increased in both MGI and BioVU, a concordant decrease effect if p-value significance decreased in both MGI and BioVU or a discordant effect if the p-value increased in significance in one cohort but decreased in the other.

#### Meta-analysis

We meta-analyzed the GWAS results from the MGI and BioVU default analysis (mean trait value, all available measures, no disease covariates). The meta-analysis was performed using *metal* by combining study-specific GWAS effect size estimates and standard errors ^25^. We computed genomic control inflation factors (*λ*_*GC*_) on a set of LD-pruned SNPs for each meta-analyzed lab.

#### GWAS Catalog Variants

We created a list of previously identified genetic associations for our analyzed lab traits using the GWAS catalog^26^ (downloaded 9/27/2017). We searched the catalog for quantitative phenotypes matching our analyzed labs using pattern matching in the DISEASE_TRAIT, MAPPED_TRAIT, and P_VALUE_TEXT columns. We searched for each lab using multiple potential string patterns, for example “AST”, “aspartate aminotransferase”, “SGOT”, and “serum glutamine oxaloacetic aminotransferase”. For purposes of replication, we limited our catalog search to studies of European cohorts performed on adults of both sexes without disease-based sampling (e.g. glucose measurements in type 2 diabetes samples) and required a reported p-value of at least 5e-8. We considered a catalog association replicated if the meta-analysis p-value for our corresponding lab was < 0.05 and the BioVU and MGI studies had the same direction of effect.

#### Definition of novelty

We report several novel lab-SNP associations reaching genome-wide significance that have not been previously reported and are not reasonably expected based on existing SNP-lab associations in similar labs and/or non-European populations. We used the following criteria to declare a lab-SNP association as a novel finding: genome-wide significance (meta-analysis p-value to be <5e-8), consistent direction of effect between MGI and BioVU and at least 1 megabase from any previously reported SNP for the given lab or a related lab in any population. Here, we define related labs as those which are commonly ordered as part of a panel of correlated tests, for example AST and ALT for liver function, and therefore likely indicate the same biological association. We report the “peak” or most significant SNP when a group of novel SNPs are in linkage disequilibrium.

#### Replication of Novel Associations

We performed a replication analysis of all novel SNP-lab associations identified in the meta-analysis using an independent cohort of BioVU samples that were made available after the original meta-analysis was performed. This replication cohort consisted of 29,043 European ancestry adult individuals with extant lab data recruited using the same procedure as the initial BioVU cohort, genotyped on the same MEGA genotyping array, and subjected to the same data QC procedure. We declared a novel SNP-lab association to be replicated if the replication p-value was <0.05 and the direction of effect was consistent with that from the meta-analysis.

## Results

We extracted all available clinical lab measurements from the electronic health records (EHRs) for genotyped samples in two academic biobank cohorts: the Michigan Genomics Initiative^16^ (MGI) at Michigan Medicine and the BioVU^15^ at Vanderbilt University. We focused on samples of European ancestry in both cohorts due to insufficient sample sizes in other ancestry groups. In total, our data consisted of 35,785,074 individual labs measurements across 5,187 distinct lab types measured in 50,743 consented MGI. The EHR-based lab measurements required extensive curation prior to genetic analysis due to the complexity of the data and the non-uniform recording between health systems. We first identified labs recorded in both health systems that could be meta-analyzed by manual matching of database names and clinical descriptors. We required at least 1,000 samples to have the lab measured in each health system and arrived at a set of 70 labs matched with high confidence (Table 1).

**Table 1:**
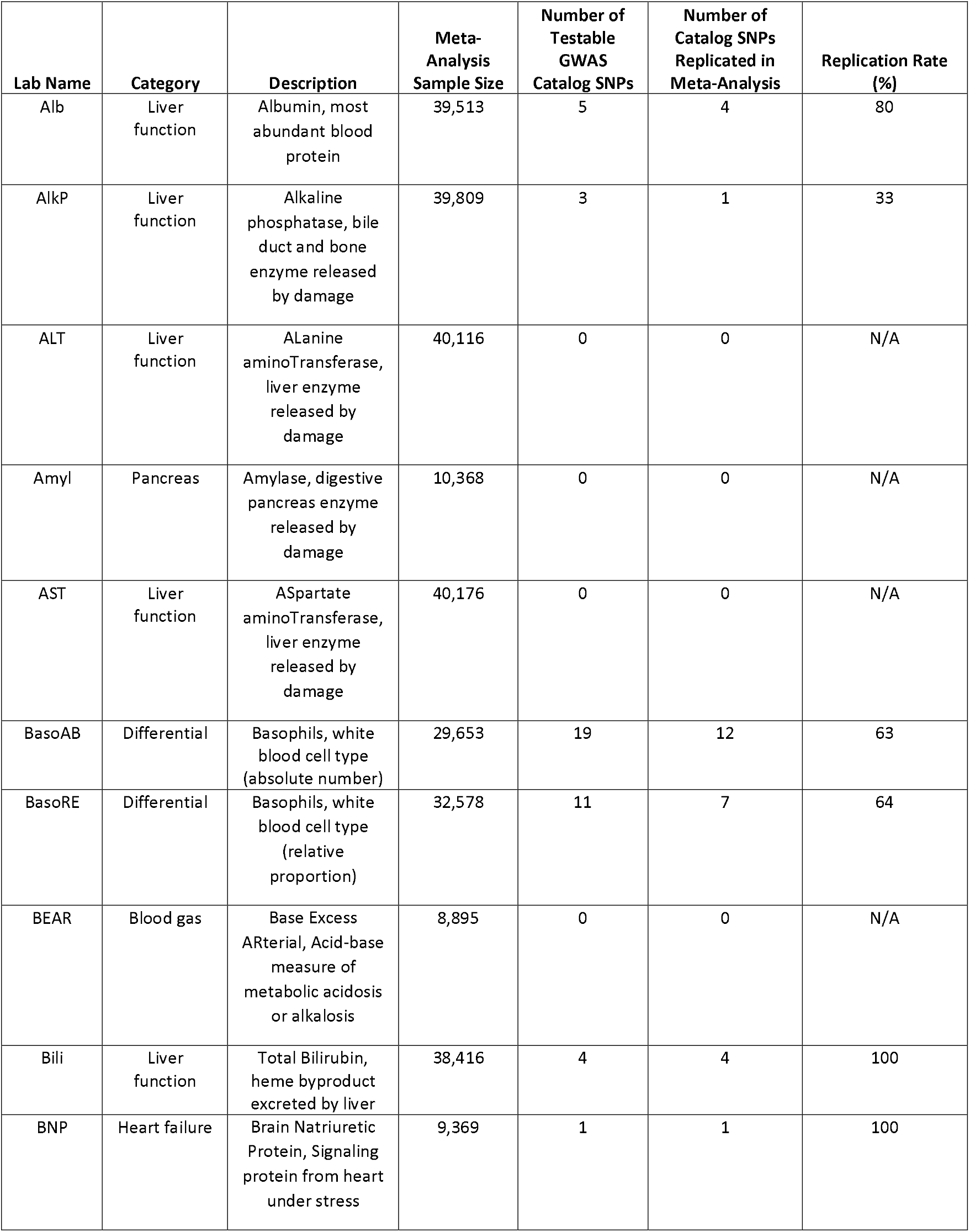
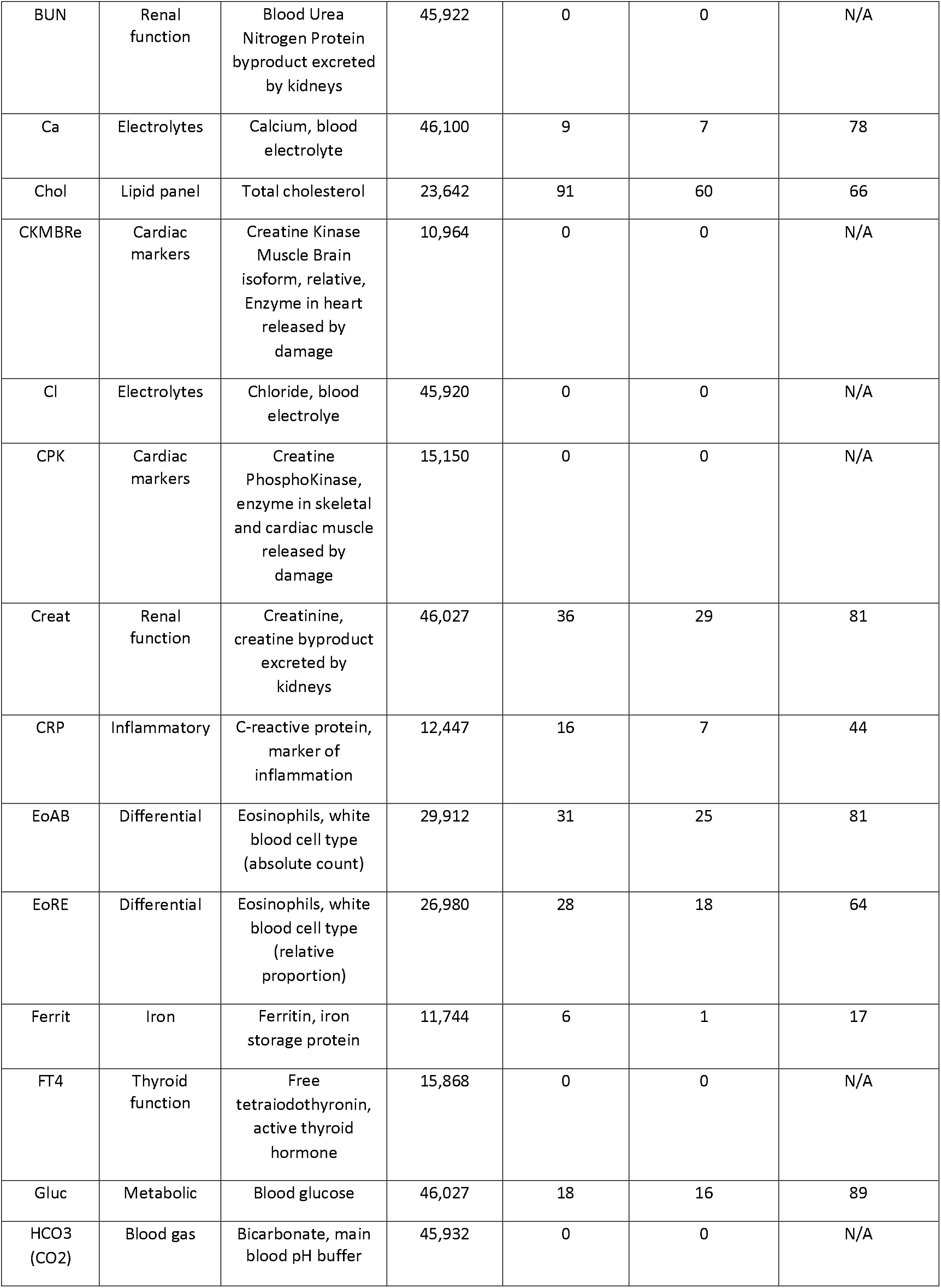
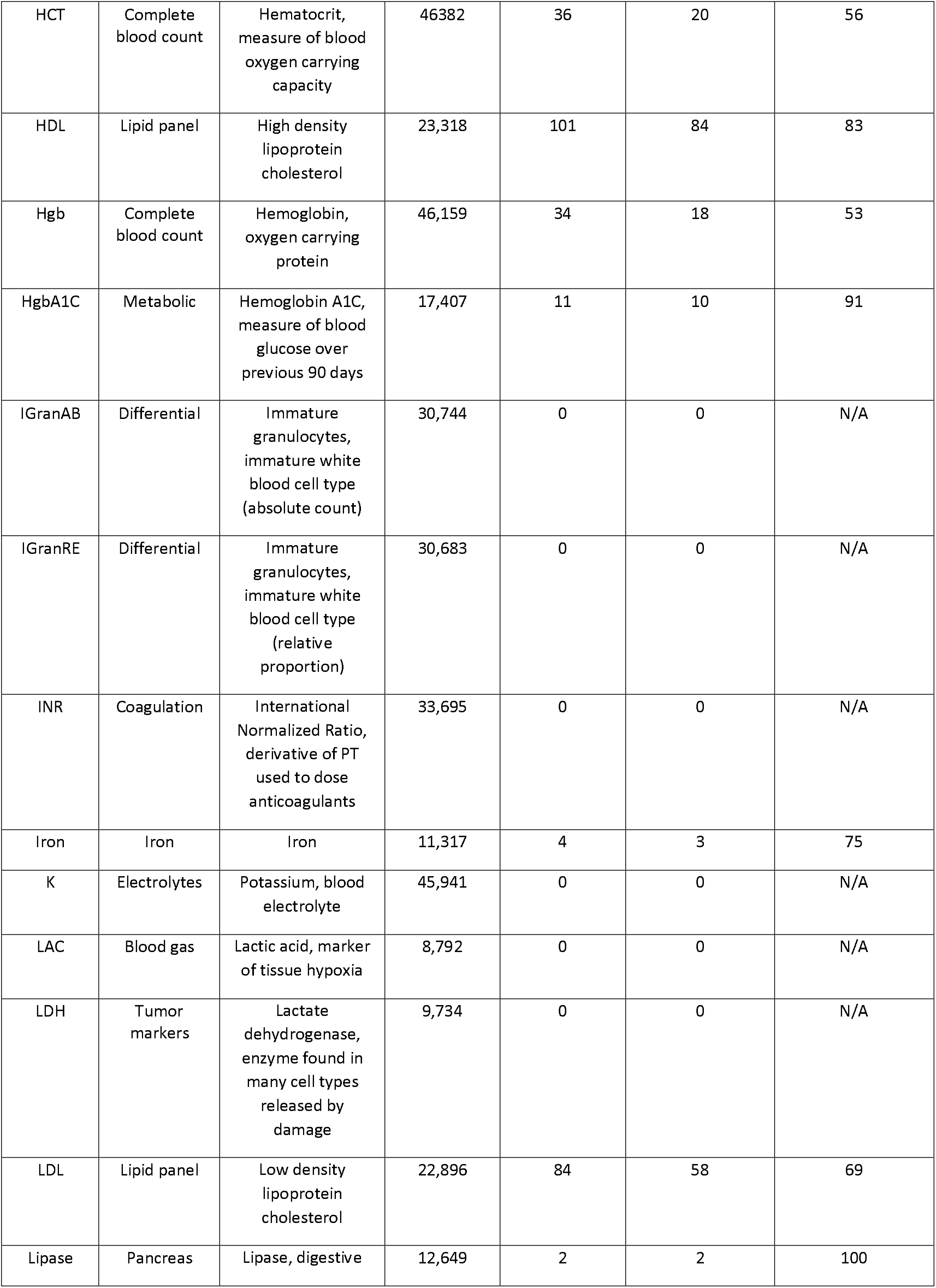
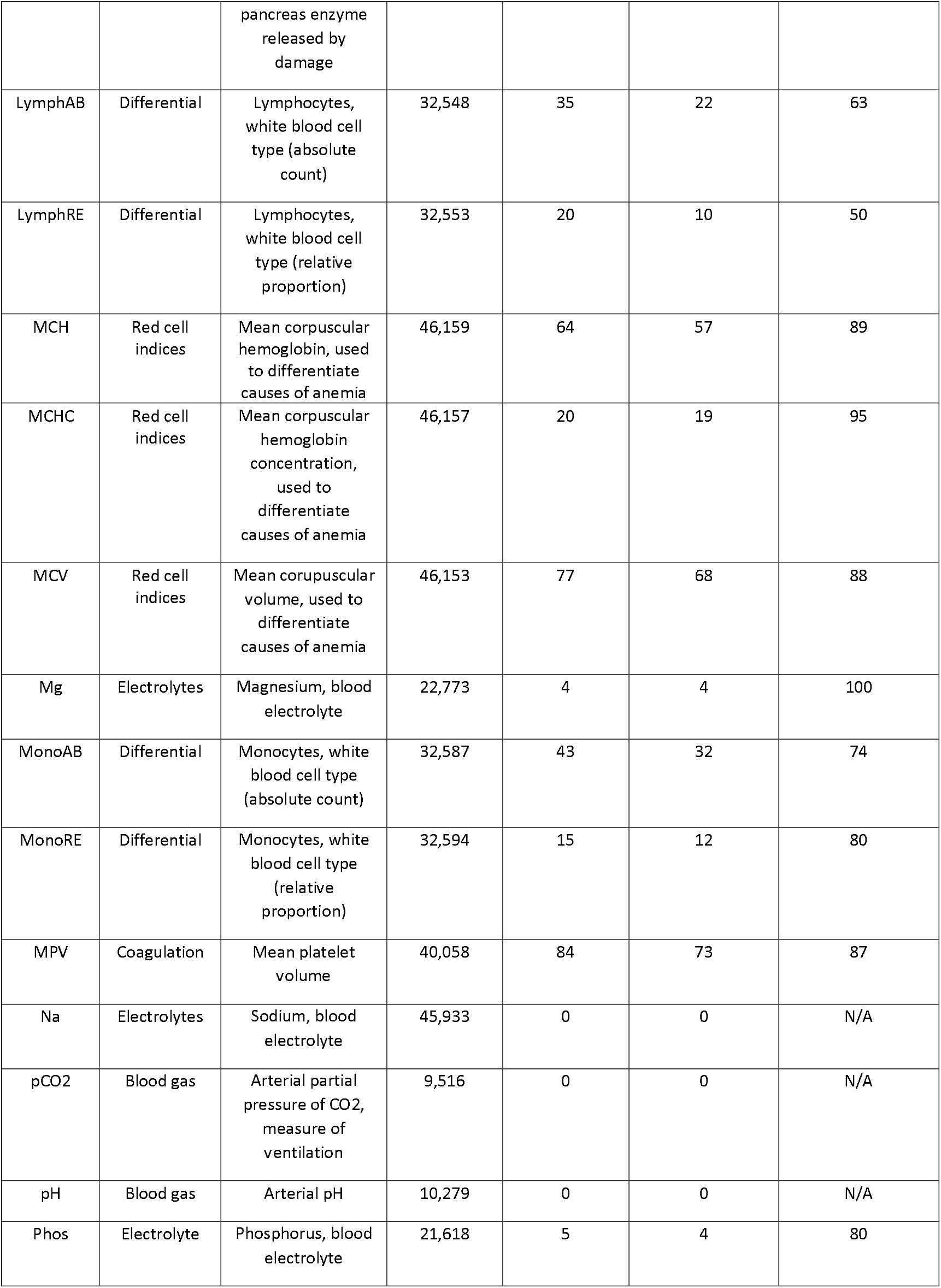
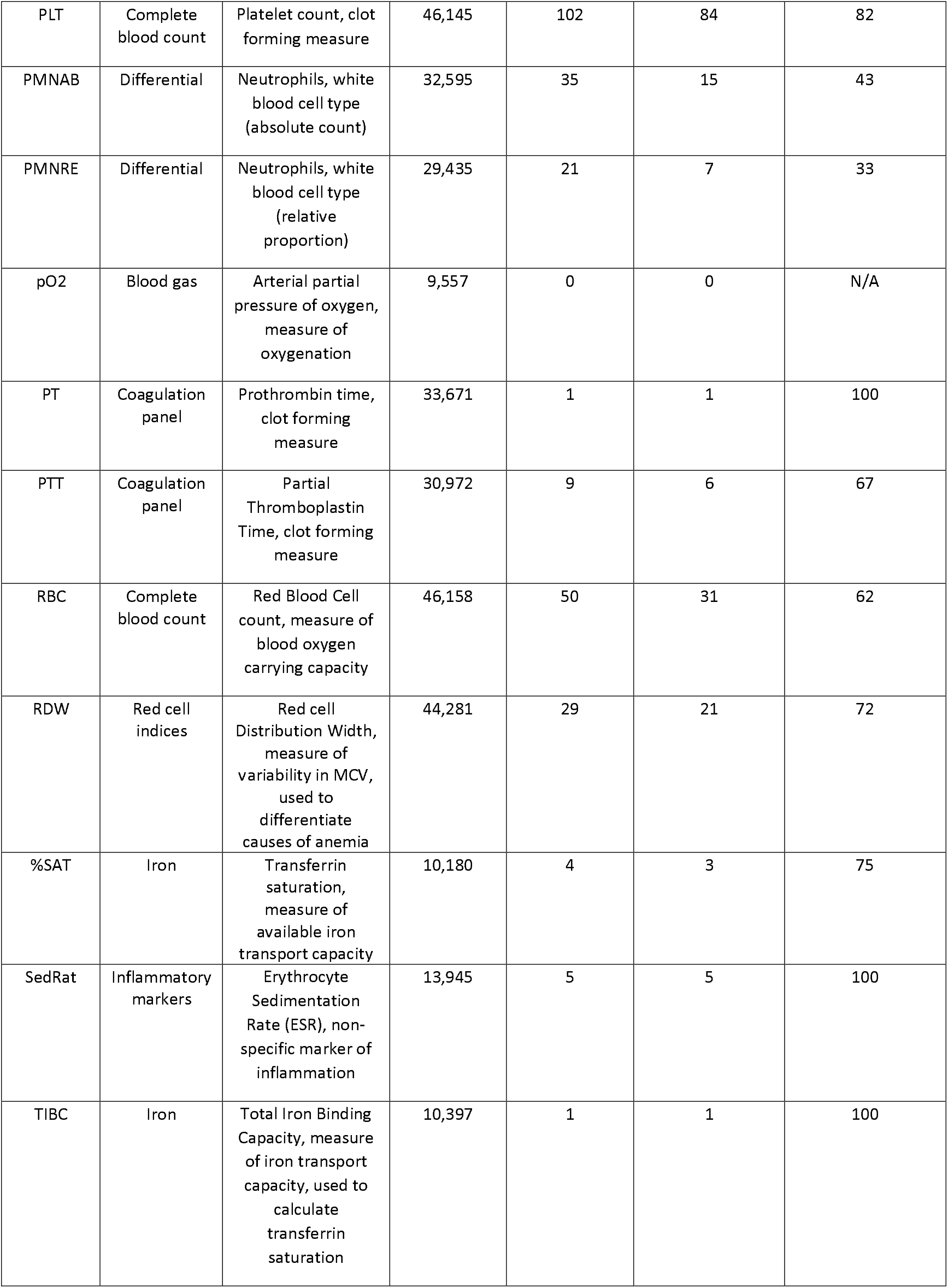
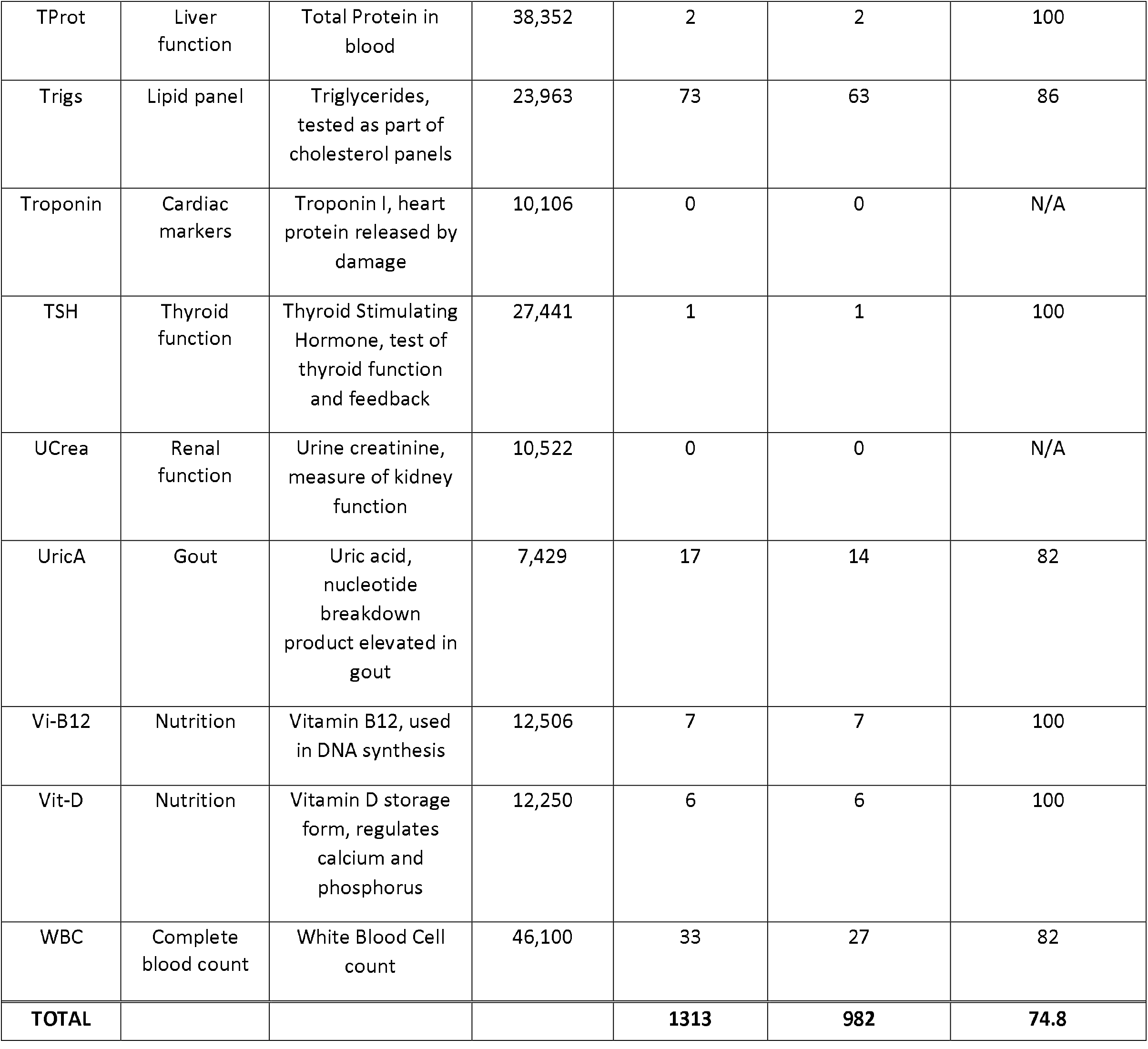
Summary of Clinical Lab measurements tested, including meta-analysis samples size, number of testable GWAS catalog SNPs, Number of replicated Catalog SNPs, Replication Rate. **Summary, labs tested, replication rate, novel findings**

Genetic analyses were performed on the set of ∼800K overlapping SNPs between the MGI imputed genotypes and the BioVU MEGA array genotypes. We identified a set of known genetic associations for our seventy matched labs based on a review of the GWAS Catalog. These catalog SNPs served as “true positive” variants to validate the data and assess various analysis strategies. We identified 1313 distinct SNP-lab associations across 48 labs that could be tested in our cohort and had previously been reported at genome-wide significance in European populations (Table 1). Many lab traits have been well studied ^27,28^ and provided many testable catalog SNPs. LDL, for example, had 84 catalog SNPs that could be directly tested in our meta-analysis. Alternatively, several labs had relatively few or no catalog SNPs, including labs for which either no variant was reported in the catalog or the catalog variants were not typed in one or more of our cohorts.

### Meta-Analysis of Labs in MGI and BioVU

The 70 EHR-derived clinical lab traits were first analyzed separately in the cohorts using the same analysis strategy: GWAS performed on inverse-normalized distribution of mean lab values across all available measures age, sex and 4 PCs included as covariates in the regression model. The combined sample size for the meta-analysis differed between labs, ranging from 7,429 for uric acid to 46,382 for hematocrit (Figure 1), reflecting the frequency with which different labs are administered in health systems. Several labs have previously been studied in much larger cohorts, including the differential panel of 10 white blood cell measures, analyzed in >170K samples in the UK BioBank^28^. However, this meta-analysis provides the largest sample size for 34 labs, including 14 clinical lab traits with no previously reported study in the GWAS catalog at the time of our analysis. Genomic control lambda values (*λ*_GC_) confirmed the meta-analyses were well-controlled^29^. The mean *λ*_GC_ across all labs was 1.035, ranging between 0.995 and 1.103. Consistent with polygenicity^30^, traits with a larger numbers of catalog variants had, on average, larger *λ*_*GC*_ values. The mean *λ*_*GC*_ for labs with zero testable catalog SNPs was 1.020. Labs with one to twenty testable Catalog SNPs had mean *λ*_*GC*_ of 1.028 and labs with greater than 20 testable Catalog SNPs had mean *λ*_*GC*_ of 1.066.

**Figure 1:**
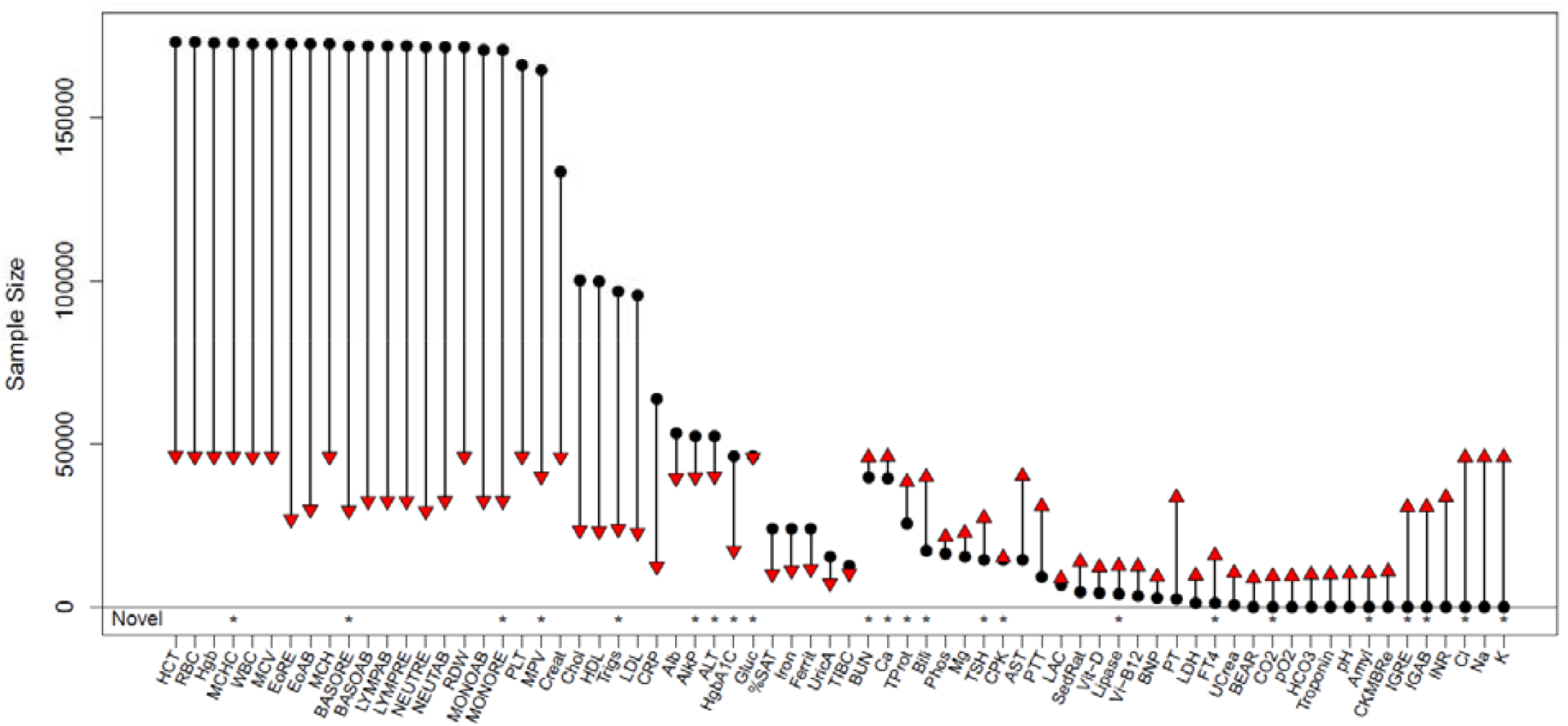
Sample sizes for 70 clinical lab traits from the meta-analysis of BioVU and MGI EHRs (red triangles) and the previous largest reported GWAS in a European cohort (black circles). Our meta-analysis provides the largest GWAS for 34 lab traits, including the first for 14. Asterisks along the bottom row indicate labs for which we identified a novel genetic association.

The complete set of meta-analysis summary statistics are viewable through an interactive *PheWeb* web browser, available at http://pheweb.sph.umich.edu/mgi-biovu-labs. This tool makes our results broadly available to the research community, allowing users to replicate their own findings or perform hypothesis-driven lookups on specific SNPs or labs of interest.

#### Replication of GWAS Catalog SNPs

We first performed a replication analysis of the 1313 GWAS catalog SNPs to validate the EHR-derived lab phenotypes. We defined a Catalog SNP as replicated if the meta-analysis p-value for the appropriate lab was <0.05 and the directions of effect were consistent between the MGI and BioVU cohorts. Overall, we replicated 982 of the GWAS catalog SNPs, giving an overall replication rate of 74.8%. Replication rates varied across the individual labs; however we did replicate at least one catalog SNP for each of the 48 traits with a testable catalog SNP (Table 1). Replication rates were high for several previously well-studied traits, including red blood cell indices (MCHC, MCH, MCV) and metabolic measures (glucose and HgbA1C) and creatinine. The lowest replication rates occurred for the differential panel of white blood cell traits (neutrophils, lymphocytes) which included catalog SNPs discovered in the much larger UKBB cohort. Interestingly, replication rates differed among the well-studied lipid panel traits. We replicated a lower percentage of catalog SNPs for LDL cholesterol and total cholesterol compared to triglycerides and HDL cholesterol.

Several factors influenced our ability to replicate individual catalog SNPs (Figure 2), each consistent with statistical power rather than adequate matching of labs as the primary limiting factor for replication. Replication increased sharply with PMID count, the number of publications reporting the association (Figure 2A). Associations reported only once in the GWAS Catalog are a mix of true yet to be replicated associations and false positives, whereas associations reported more than once have already been replicated and are likely real. We replicated 70% (699 of 1000) of associations reported only a single time. That rate increased to 77% (196 of 256) for associations reported twice, 91% for associations reported three times and nearly 100% (56 of 57) for associations reported four or more times. Importantly, this analysis provides the first replication for 699 previously reported SNP-lab associations, increasing the likelihood that these are true genotype-phenotype associations (Supplementary Table).

**Figure 2:**
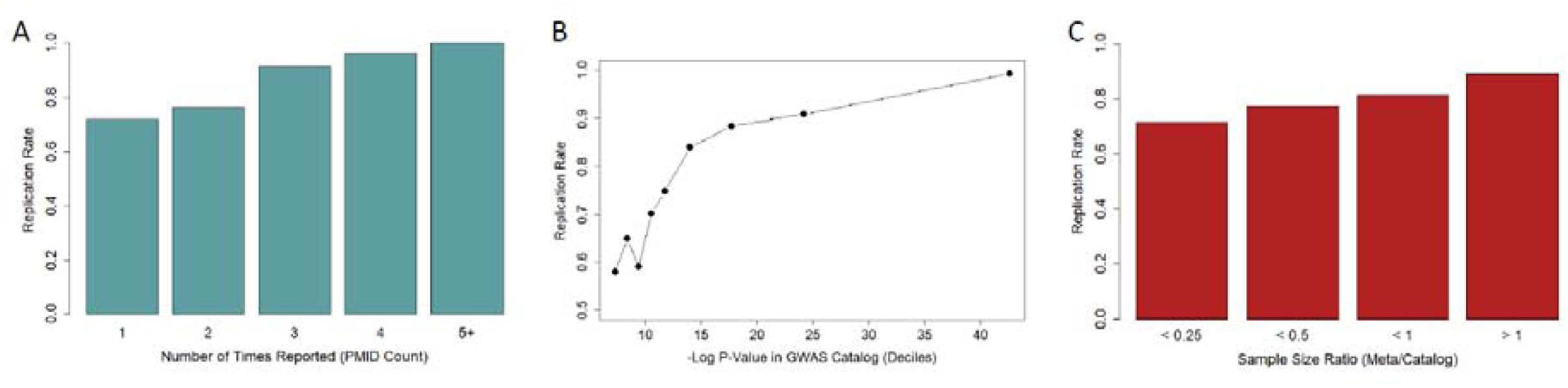
Replication rates for GWAS catalog SNPs of clinical labs increased with (A) the number of times an association was reported in the GWAS catalog, (B) the most significant p-value previously reported for the association, and (C) the ratio of sample size in our meta-analysis to that of the previous largest study.

Replication rate was also dependent on both the best previously reported p-value for the association and the sample size of the study reporting the association (Figure 2B & 2C). Our replication rate was lowest, between 55%-65%, for associations whose best reported p-value was just above genome-wide significance of 5e-8 but increased sharply thereafter. We replicated ∼85% of catalog SNPs with best reported p-value <1e-15 and over 90% of catalog SNPs with best p-value <1e-20. Replication rate increased with the relative size of our meta-analysis compared to the largest reported study. We replicated approximately 90% of catalog SNPs for which our meta-analysis was at least as large as prior studies reporting the association.

#### Novel SNP-Lab Associations

We next searched for novel associations across the 70 meta-analyzed lab traits. We defined an association to be novel if it attained genome-wide significance (meta-analysis p < 5e-8), had consistent directions of effect in MGI and BioVU, was >1 megabase away from a GWAS catalog SNP for the lab, and had not been reported in any population or in a related lab (e.g. LDL cholesterol for total cholesterol). In total, we identified 264 SNP-lab pairs satisfying our definition of novelty. Based on visual inspection, the novel SNPs corresponded to 31 distinct peaks for which we report the lead SNP having the strongest association signal at each peak (Table 2).

**Table 2:** Summary of Novel findings.

| Lab | SNP | Chr:Pos | Allele 1 | Allele 2 | MGI-BioVU Meta-Analysis |  |  | BioVU Replication Cohort |  |  | Replicated |
| --- | --- | --- | --- | --- | --- | --- | --- | --- | --- | --- | --- |
|  |  |  |  |  | N | Beta | P-Value | N | Beta | P-Value |  |
| AlkP | rs3843738 | 17:43739194 | A | G | 39,809 | 0.04 | 2.51E-08 | 22,920 | 0.01 | 3.58E-01 | No |
| AlkP | rs73004933 | 19:19675696 | T | C | 39,809 | 0.08 | 4.47E-09 | 22,730 | 0.05 | 7.14E-03 | Yes |
| ALT | rs112574791 | 8:145730221 | A | G | 40,116 | 0.18 | 3.02E-08 | 23,007 | 0.15 | 5.80E-04 | Yes |
| Amyl | rs1930212 | 1:104324819 | A | G | 10,368 | -0.25 | 1.48E-45 | 3,573 | -0.18 | 4.69E-09 | Yes |
| Amyl | rs8051363 | 16:75255217 | A | G | 10,368 | 0.10 | 1.07E-10 | 3,564 | 0.09 | 4.51E-04 | Yes |
| BasoRE | rs386785158 | 15:70744437 | T | C | 29,653 | 0.06 | 7.94E-13 | 16,191 | 0.04 | 2.10E-04 | Yes |
| Bili | rs855791 | 22:37462936 | A | G | 39,890 | 0.04 | 2.34E-08 | 22,918 | 0.04 | 1.00E-05 | Yes |
| BUN | rs10516957 | 4:95949206 | T | C | 45,922 | -0.06 | 1.35E-08 | 25,245 | 0.01 | 6.11E-01 | No |
| Ca | rs6727384 | 2:97400324 | A | G | 46,100 | -0.04 | 5.13E-10 | 25,200 | -0.05 | 2.06E-07 | Yes |
| Ca | rs2839899 | 9:80350999 | A | G | 46,100 | 0.04 | 6.76E-09 | 25,194 | 0.03 | 9.47E-03 | Yes |
| Cl | rs1030025 | 2:103105611 | A | T | 45,920 | 0.05 | 4.68E-10 | 25,204 | 0.02 | 9.16E-02 | No |
| FT4 | rs10122824 | 9:139109861 | T | G | 15,868 | 0.07 | 1.00E-09 | 9,721 | 0.07 | 7.28E-07 | Yes |
| Glucose | rs7607980 | 2:165551201 | T | C | 46,027 | -0.05 | 4.27E-09 | 25,312 | -0.04 | 2.09E-03 | Yes |
| Glucose | rs896854 | 8:95960511 | T | C | 46,027 | -0.04 | 1.55E-09 | 25,311 | 0.01 | 3.64E-01 | No |
| Glucose | rs9273364 | 6:32626302 | T | G | 46,027 | 0.05 | 2.63E-11 | 24,801 | 0.05 | 3.10E-06 | Yes |
| HgbA1C | rs3130628 | 6:31609272 | T | C | 17,407 | -0.08 | 1.23E-08 | 7,340 | 0.03 | 3.79E-02 | No |
| HCO3 (CO2) | rs1799913 | 11:18047255 | T | G | 45,932 | -0.04 | 5.89E-09 | 25,219 | -0.04 | 7.82E-07 | Yes |
| HCO3 (CO2) | rs77375846 | 2:103155075 | T | C | 45,932 | -0.10 | 9.33E-25 | 25,217 | -0.06 | 2.78E-05 | Yes |
| IGranRE | rs13284665 | 9:131513370 | A | G | 30,683 | 0.22 | 6.61E-74 | QC Fail | N/A | N/A | No |
| IGranAB | rs13284665 | 9:131513370 | A | G | 30,744 | 0.13 | 6.76E-35 | QC Fail | N/A | N/A | No |
| K | rs10039139 | 5:137164863 | T | G | 45,941 | 0.07 | 8.32E-16 | 25,211 | 0.06 | 1.83E-06 | Yes |
| Lipase | rs9377343 | 6:96512220 | A | G | 12,649 | -0.10 | 4.79E-14 | 5,564 | -0.08 | 3.60E-05 | Yes |
| Lipase | rs8051363 | 16:75255217 | A | G | 12,649 | 0.13 | 2.00E-20 | 5,549 | 0.07 | 8.39E-04 | Yes |
| MCHC | rs12352830 | 9:80041132 | C | G | 46,157 | -0.04 | 4.37E-08 | 26,243 | -0.04 | 5.77E-05 | Yes |
| MonoRE | rs117358683 | 12:44145965 | A | G | 32,594 | -0.23 | 2.69E-08 | 16,185 | 0.04 | 4.07E-01 | No |
| MPV | rs11212635 | 11:108310702 | A | T | 40,058 | 0.04 | 9.55E-09 | 17,333 | -0.01 | 3.68E-01 | No |
| TProt | rs8022180 | 14:103263020 | A | G | 38,352 | 0.04 | 7.24E-10 | 19,665 | 0.03 | 2.63E-03 | Yes |
| Trigs | rs6847598 | 4:76750356 | T | C | 23,963 | -0.05 | 1.58E-08 | 12,526 | -0.03 | 1.48E-02 | Yes |
| TSH | rs12590163 | 14:105223525 | T | C | 27,441 | -0.05 | 4.68E-08 | 17,042 | -0.04 | 6.76E-04 | Yes |
| TSH | rs310766 | 3:12233482 | A | G | 27,441 | -0.06 | 1.66E-08 | 17,079 | -0.05 | 1.42E-05 | Yes |
| TSH | rs9275141 | 6:32651117 | T | G | 27,441 | 0.05 | 3.47E-09 | 17,054 | 0.04 | 8.64E-04 | Yes |

We performed a replication analysis of the 31 novel lead SNPs using an independent cohort of 29,043 BioVU patients that became available after the initiation of our primary analysis. We considered the novel association to be replicated if the lead SNP had p-value < 0.05 in the replication cohort and the direction of effect was consistent with our initial meta-analysis (Table 2). One SNP that was potentially novel for both immature granulocytes measures failed QC filtering in the replication cohort and could not be tested for replication. In total, we replicated 22 of the 31 (71%) novel associations. Among the 24 replicated novel SNPs are the first associations for amylase (Amyl) and bicarbonate (CO2). We identified and replicated additional associations for alanine aminotransferase (ALT), alkaline phosphate (AlkP), Relative count of basophils (BasoR), total bilirubin (Bili), calcium (Ca), creatinine phosphokinase (CPK), glucose (gluc), mean corpuscular hemoglobin concentration (MCHC), lipase, and thyroid stimulating hormone (TSH).

Several of our novel findings have biological or existing evidence that support the association. Three of the associations have recently been identified for the same lab in non-European cohorts. rs855791, a missense variant in *TMPRSS6* (transmembrane serine protease 6), and rs8022180, an intronic variant in *TRAF3*, were shown to be associated with bilirubin and serum total protein level, respectively, in a Japanese population^10^. rs112574791 is in the glutamic--pyruvic transaminase gene *GPT*, a gene associated with alanine aminotransferase levels in the Korea Biobank^31^. Our results confirm these prior findings and suggest a cross-ethnic effect in European populations.

The intronic variant rs8051363 in *CTRB1* was associated with both amylase and lipase, clinical assays of pancreas function used to diagnose pancreatitis. While the SNP itself has previously been linked to blood protein measurements^32^, the *CTRB1* gene encodes chymotrypsin, a component of digestive enzyme secreted by the pancreas, and was previously shown to be associated with alcoholic chronic pancreatitis^33^. A second novel SNP for lipase, rs9377343 is an intronic variant in *FUT9*, a gene that showed association with diabetic neuropathy in a trans-ethnic meta-analysis^34^.

The amylase-associated SNP rs1930212 resides near three amylase genes (*AMY2B, AMY2A* and *AMY1*) on chromosome 1, each of which encodes enzymes that digest starch into sugar^35^. Copy number variation for amylase genes is hypothesized to have been subject to selective sweeps corresponding to starch content in human diets^36^. The rs1930212 SNP tags a known deletion of *AMY2A*, a pancreatic amylase enzyme, most common in populations historically lacking starch rich diets^36^.

One of our novel results for calcium, rs2839899, is an intronic variant in *GNAQ* (G protein subunit alpha q), a signaling protein involved in response to various hormones. Variation in *GNAQ* is associated with Sturge-Weber syndrome^37^, a hereditary vascular malformation syndrome which can lead to deposits of calcium (calcification) in the brain.

Three SNPs showed associations with glucose. rs7607980 is a missense variant in *COBLL1* previously linked to fasting blood insulin and Type 2 diabetes^38–40^. rs9273364 is located near HLA-DQB1-AS1, a gene associated with T2D^41^. And, although it did not replicate in our analysis, rs896854, a variant mapping to both *NDUFAF6* and *TP53INP1*, has recent associations with T2D^42^ and eosinophil count^43^ among UK biobank participants.

We note that several associations occurred within the HLA region on chromosome 6, notably for glucose, hemoglobin A1C, and TSH. These variants are likely segregating with HLA types, which are strongly associated with various autoimmune diseases including diabetes and autoimmune thyroiditis, which have strong effects in these particular labs.

#### Genetic Correlation of Clinical Labs

We computed the genetic correlation between pairs of labs to learn about shared genetic basis of these traits. We computed the correlations using LD score regression, restricting analysis to the 50 lab traits with heritability of at least 7%. The heatmap in Figure 3 shows the correlation structure of the labs, noting only correlations with p<0.05. We observe several clusters with strong positive correlations among lab traits of similar function. The liver enzymes alanine aminotransferase (ALT) and aspartate aminotransferase (AST) were strongly correlated, as were the measures of renal function Blood Urea Nitrogen (BUN) and creatinine (Creat). Prothrombin time (PT), a measure of clot formation time and a derivative measure International Normalized Ratio (INR) were, not surprisingly, positively correlated. Interestingly, INR was also positively correlated with vitamin D. While vitamin K is known to be required for the formation of prothrombin, this represents a novel association. Their correlation suggests covariance in nutrition or nutrient absorption.

**Figure 3:**
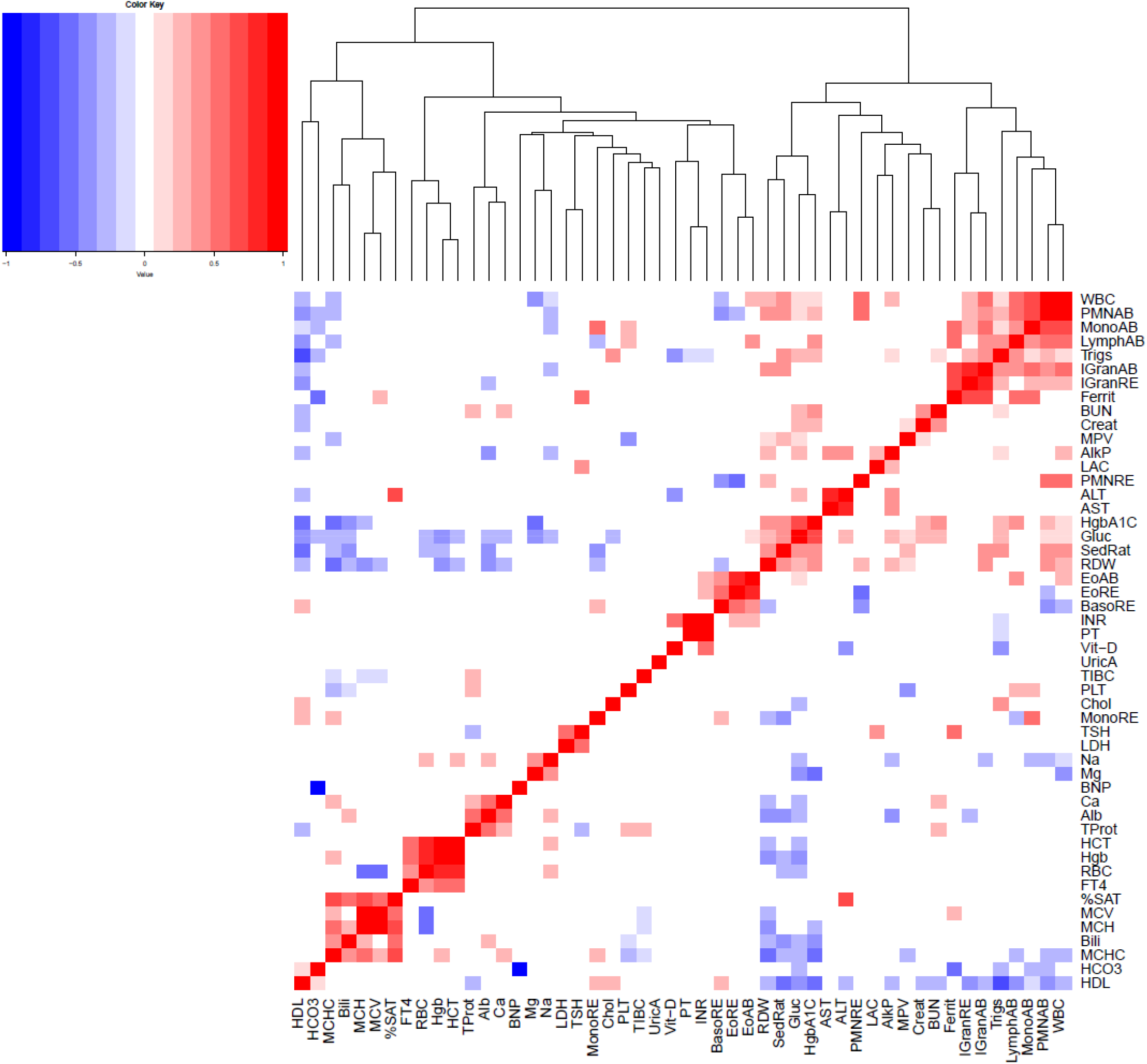
Pairwise genetic correlation of clinical lab traits. We restricted to labs with heritability of at least 7%. Squares are colored only for correlations having a p-value <0.05 for the null hypothesis of correlation equal to zero.

A prominent cluster of labs (top right corner of the heatmap) contains primarily white blood cell traits including measures of immature granulocytes, lymphocytes, monocytes and neutrophils. The immature granulocytes also showed a strong correlation with ferritin (ferrit). Ferritin is an iron storage protein as well as an acute phase protein. In severe acute inflammation, ferritin and immature granulocytes could both be elevated.

HgbA1C and glucose were, not surprisingly, strongly correlated. More interestingly, they also clustered with Red cell Distribution Width (RDW) and Erythrocyte Sedimentation Rate (SedRat). This cluster of labs showed negative associations with high density lipoprotein (HDL), mean cell hemoglobin concentration (MCHC), and mean cell hemoglobin (MCH). This supports a pathophysiology where the metabolic syndrome (obesity, elevated glucose, low HDL) is linked by complex mechanisms to persistent low-level inflammation (elevated SedRat), and anemia of chronic disease (elevated RDW, low MCH, low MCHC).

We identified a cluster of the red cell indices – mean cell hemoglobin concentration (MCHC), mean cell hemoglobin (MCH), and mean cell volume (MCV) – with total bilirubin (Bili) and transferrin saturation (%SAT). This reflects the biology of hemoglobin – iron is carried to red cell precursors by transferrin and incorporated into heme and thence hemoglobin, red cells are filled with hemoglobin, and at the end of a red cell lifecycle, heme is broken down into bilirubin.

Additional clusters include (1) calcium (Ca), albumin (Alb) and total protein in blood (TProt), (2) thyroid stimulating hormone (TSH) and lactate dehydrogenase (LDH), and (3) hematocrit (HCT), red blood cell count (RBC) and hemoglobin (Hgb) with free tetraiodothyronine (FT4). These causes for these are correlations are not immediately clear and may suggest new biology for future study.

### Analytic strategies for EHR-derived lab traits

To understand the effect of various analytic choices on downstream analysis, we performed parallel GWAS analyses in the MGI and BioVU cohorts in which we perturbed one of the analytic steps from our original analysis: the per-sample statistic used to summarize longitudinal lab measurements and the inclusion of covariates for underlying comorbid health conditions. We performed these analyses on the 22 lab traits for which there were least 20 testable GWAS catalog SNPs, using the catalog SNPs to interpret the effect of each analytic strategy on true risk variants. For a fixed lab and cohort, we quantified the change in p-value for each SNP using Δ_*p*_, the −log_10_ fold change in p-value for an alternative analysis versus the default analysis (see Methods). A positive value of Δ_*p*_ indicates a SNP that increases in significance (smaller p-value) when the alternate summary statistic. A negative value of Δ_*p*_ indicates a decrease in significance for the alternate analysis. Scatterplots of Δ_*p*_ computed in MGI and BioVU summarize the magnitude and consistency of change in p-value significance between the cohorts (Figure 4 for an example, Supplementary Material).

**Figure 4:**
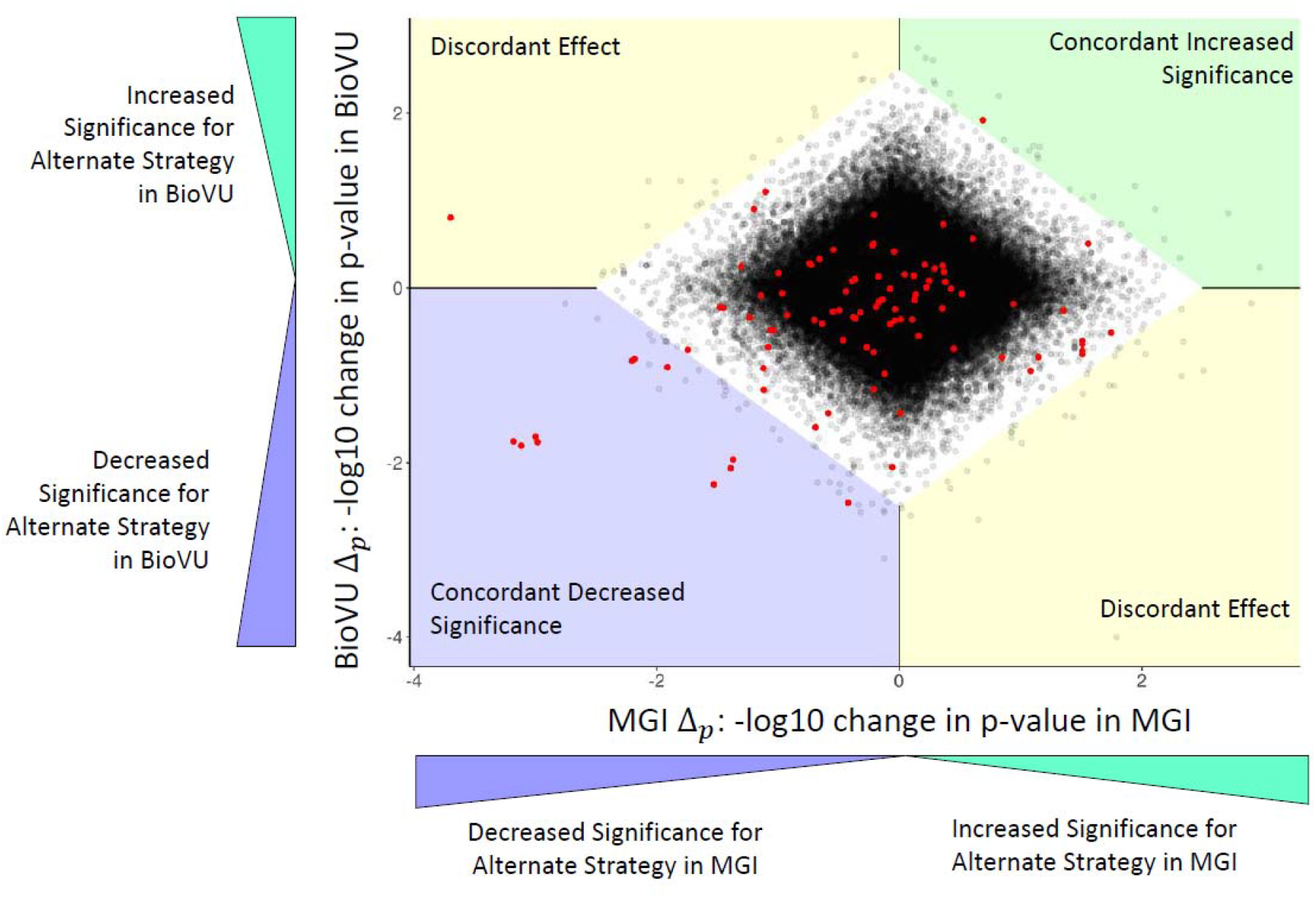
Scatterplot of Δ_p_ in MGI and BioVU when using the first available measure rather than the mean measurement in a GWAS of Cholesterol level. Δ_p_ is the −log fold change in p-value at a SNP for using an alternate analysis, in this case the first available lab measurement. Each dot is a SNP, with red dots indicating GWAS catalog SNPs for the specific lab trait. The white diamond contains 99.9% of SNPs and is used to identify SNPs with the largest changes in p-value due to the alternate analysis. SNPs outside the bounding diamond in the top right (green) quadrant show a concordant increase in significance in both MGI and BioVU, that is, SNPs for which the alternative strategy increases significance in both cohorts. Conversely, SNPs in the bottom left (blue) quadrant show a concordant decrease in significance in both MGI and BioVU. SNPs in either the top left or bottom right (yellow) quadrants have a discordant effect, indicating a large increase in p-value in one cohort but a large decrease in p-value in the second cohort. In this example, one catalog SNP showed a concordant increase in significance when using the first available lab measure, 11 catalog SNPs had a concordant decrease in significance and one SNP had discordant effects. The complete set of scatterplots for each analyzed lab and alternative analysis strategy (summary statistic and comorbidity model) are included in the Supplementary Material. Tables 3 and 4 summarize the movement of catalog SNPs for each lab and analysis strategy.

#### Summary statistic

Patients in EHRs often have multiple measurements for the same lab test taken over many years of treatment. These measurements are typically summarized into a single numeric value that is used as the phenotype in a GWAS, with the mean being a common choice for the summary statistic. We hypothesized that alternate summary statistics could result in more powerful genetic analyses. For example, the median is less sensitive to individual outlier lab measurements making it more robust against data recording errors or extreme true measurements. Alternatively, the maximum or first available lab measurement for an individual patient could mitigate the effects of prescription drugs for modifiable lab traits.

Overall, 13.3% of testable catalog SNPs showed a major change in significance when using the median as opposed to mean value for the summary statistic (Table 3). The median rarely resulted in a consistent improvement for both MGI and BioVU. Only 0.4% of catalog SNPs had concordant increased effect compared to 7.6% with concordant decreasing effect and 5.2% with a discordant effect. Creatinine was the sole lab for which using median lab value had a greater number of catalog SNPs with concordant increased significance than catalog SNPs with concordant decreased significance. Even here the effect was small, only two of the 36 catalog SNPs had a concordant increase in significance.

**Table 3:** Classification of catalog SNPs for alternate summary statistics.

|  |  | Median Measurement |  |  | First Available Measurement |  |  | Maximum Measurement |  |  |
| --- | --- | --- | --- | --- | --- | --- | --- | --- | --- | --- |
| Lab | Testable Catalog SNPs | Concordant Increased Significance | Concordant Decreased Significance | Discordant Effect | Concordant Increased Significance | Concordant Decreased Significance | Discordant Effect | Concordant Increased Significance | Concordant Decreased Significance | Discordant Effect |
| Chol | 91 | 0 | 12 | 0 | 1 | 11 | 1 | 2 | 4 | 6 |
| Create | 36 | 2 | 0 | 0 | 2 | 2 | 1 | 0 | 8 | 1 |
| EoAB | 31 | 0 | 6 | 0 | 0 | 9 | 0 | 0 | 2 | 1 |
| EoRE | 28 | 0 | 1 | 0 | 0 | 4 | 0 | 0 | 1 | 1 |
| HCT | 36 | 0 | 0 | 0 | 4 | 0 | 1 | 15 | 0 | 1 |
| HDL | 101 | 0 | 6 | 3 | 0 | 15 | 1 | 0 | 27 | 5 |
| Hgb | 34 | 0 | 0 | 0 | 5 | 0 | 0 | 12 | 0 | 0 |
| LDL | 84 | 0 | 9 | 1 | 0 | 9 | 4 | 2 | 2 | 6 |
| LymphAB | 35 | 0 | 0 | 0 | 0 | 3 | 1 | 5 | 1 | 2 |
| LymphRE | 20 | 0 | 0 | 0 | 0 | 0 | 0 | 0 | 0 | 0 |
| MCHC | 20 | 0 | 1 | 0 | 2 | 5 | 3 | 2 | 5 | 1 |
| MCH | 64 | 0 | 16 | 27 | 0 | 33 | 8 | 0 | 33 | 7 |
| MCV | 77 | 1 | 5 | 7 | 0 | 19 | 13 | 0 | 30 | 6 |
| MonoAB | 43 | 2 | 3 | 0 | 0 | 9 | 0 | 0 | 13 | 1 |
| MPV | 84 | 0 | 11 | 9 | 0 | 39 | 9 | 5 | 20 | 17 |
| PLT | 102 | 0 | 0 | 1 | 7 | 7 | 1 | 0 | 19 | 5 |
| PMNAB | 35 | 0 | 0 | 0 | 0 | 2 | 1 | 0 | 3 | 1 |
| PMNRE | 21 | 0 | 0 | 0 | 0 | 0 | 0 | 0 | 0 | 1 |
| RBC | 50 | 0 | 4 | 4 | 13 | 0 | 1 | 21 | 0 | 0 |
| RDW | 29 | 0 | 1 | 2 | 0 | 1 | 4 | 0 | 7 | 0 |
| Trigs | 73 | 0 | 7 | 0 | 1 | 15 | 1 | 0 | 22 | 0 |
| WBC | 33 | 0 | 4 | 5 | 0 | 7 | 1 | 0 | 9 | 0 |
| Total | 1127 | 5<br>(0.4%) | 86<br>(7.6%) | 59<br>(5.2%) | 35<br>(3.1%) | 190<br>(16.9%) | 51<br>(4.5%) | 64<br>(5.6%) | 206<br>(18.3%) | 62<br>(5.5%) |

**Table 4:**
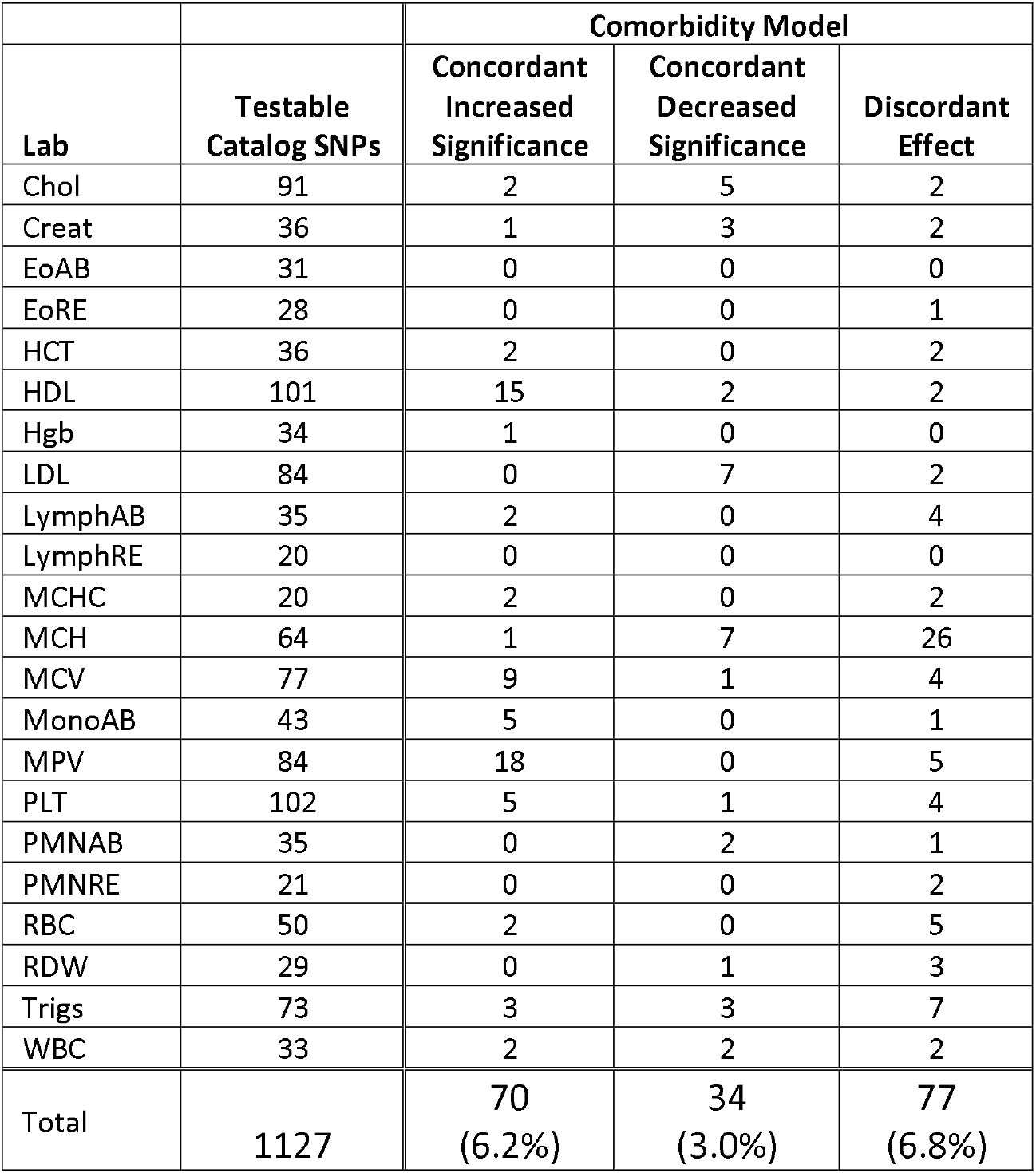
Classification of catalog SNPs for the comorbidity model, which includes covariates for various lab-altering diseases.

In comparison, the first available measurement and the maximum measurement had a greater impact on association p-values for catalog SNPs. In both cases, the alternate summary statistic was most likely to cause a concordant decrease in significance. Using the first available measurement resulted in concordant increase for only 3.1% of catalog SNPs, whereas 16.9% of catalog SNPs had a concordant decrease and 4.5% had discordant changes in significance. Using the maximum available measure had similar performance (5.6% concordant increase, 18.3% concordant decrease, 5.5% discordant).

Despite an overall trend of reducing significance of known risk variants, several related labs for blood oxygen carrying capacity did benefit from using the first available or maximum measurements. Red blood cell count (RBC), hematocrit (HCT) and hemoglobin (Hgb) each showed concordant increase in significance for several of their respective catalog SNPs without negatively impacting remaining catalog SNPs. This likely reflects red cell biology. Conditions that decrease oxygen carrying capacity, such as blood loss or iron deficiency are far more common than those that increase it, polycythemia vera or severe obstructive sleep apnea, for example. Thus, maximum measurement of an individual’s oxygen carrying capacity more likely represents the genetically determined set point.

#### Controlling for disease

The real-world health system cohorts feature a heterogeneous collection of disease comorbidities that can impact lab measurements in complex ways. One strategy of accounting for individual-level patient health is the inclusion of lab-mediating diseases as covariates in a regression model, a strategy employed by a prior GWAS of lab values in the JapanBiobank^10^. To test the efficaciousness of simultaneously controlling for the whole “kitchen sink” of diseases, we performed a GWAS using a comorbidity model which included binary covariates for 42 diseases with the potential to alter lab values.

The comorbidity model produced the largest proportion of catalog SNPs (6.2%) with concordant increased significance in MGI and BioVU among the alternate analysis strategies considered. Despite this, a roughly equal number of catalog SNPs had discordant effects (6.8%) between the two cohorts.

The clearest example of a substantial and consistent effect on catalog SNPs between MGI and BioVU was for HDL and Mean platelet volume (MPV). In contrast to the improvement for many catalog SNPs for HDL, LDL had interestingly no catalog SNPs with concordant increase in significance and seven catalog SNPs with concordant decrease.

## Discussion

This study represents the first cross-health system study of EHR-derived lab traits at large scale. We performed whole genome analysis of 70 lab traits and have made these association results easily accessible to the research community. Thoroughly dissecting each lab-SNP combination is a daunting task. Here, we focused on replication of GWAS catalog variants to validate our data and highlighted novel genetic associations. We anticipate that our full results, including those which do not reach genome-wide significance will be useful in replicating future novel results, in studies which synthesize findings across multiple SNPs, or in hypothesis-driven studies which require less stringent thresholds.

Our study serves as a proof-of-principle for performing cross-health-system genetic analysis of EHR-derived lab values. The high replication rate for known GWAS variants proves that EHR lab values can be well-matched between discordant health systems. Moreover, the replication analysis showed that EHR measurements, taken during real-life medical interactions, accurately reflect those taken under more idealized experimental conditions of previous GWAS. This also implies that mechanisms underlying variation in labs in healthy populations also act in a mixed population of patients with disease, strengthening their clinical relevance. By comparing various analytic strategies, we show that there is no optimal strategy that holds across all labs. In fact, we observed many instances in which the alternate analysis simultaneously increased significance for some risk variants and decreased significance for others. Thus, even within a given lab an optimal strategy might not exist. A potential area of future research would be determining if multiple versions of a lab trait can be combined into an omnibus test that simultaneously increases power across all risk variants. We encourage researchers to use our results across the various analysis strategies to guide decisions about how best to analyze their traits of interest.

The primary strength of our study was the access to two independent biobank cohorts. Using two cohorts provides an obvious increase in sample size and power over analyzing and reporting on each cohort separately. In addition, the two-cohort design adds a built-in internal consistency check to our results by requiring effect sizes to be in the same direction in both cohorts. This additional requirement reduced the potential for unknown biases in the health system cohorts to create spurious results when replicating GWAS catalog SNPs or novel association discovery. Further, the independent cohorts provided the means to rigorously examine analytic strategies for biobank cohorts. The heterogeneous nature of EHRs and ascertainment schemes magnify the need to replicate findings. Our mirrored analyses revealed provided the means to confirm consistent effects for analytic strategies in independent cohorts. A single cohort methodologic study could produce recommendations that are over fitted to one specific context. Use of multiple sites increases the generalizability of our recommendations. This study was further strengthened by the fortuitous availability of an independent tranche of BioVU samples that provided a replication cohort for the novel findings of our primary meta-analysis.

Our study has implications for the design and analysis of similar studies in the future. Matching and analyzing labs between health systems is difficult and requires substantial content knowledge. This study benefited from a multi-disciplinary team consisting of clinical experts to lead the categorization of the raw lab data extracts and statistical geneticists to guide analytic strategies. We leaned heavily on GWAS catalog SNPs to serve as positive controls. When possible, researchers should incorporate an explicit replication step to validate lab data quality prior to testing novel hypotheses. Summarizing the longitudinal individual-level lab measurements using the simple definition of mean value taken on all available measures proved relatively robust across labs but was by no means optimal in all scenarios. Future studies can benefit from considering the specific lab trait being evaluated. The consistency of analytic strategies is important for using EHR-based GWAS as replication datasets. Attention must be paid to the differences in preparation of EHR phenotypes, particularly for longitudinal lab measurements. Failure to replicate a finding can be due to actual lack of a true effect but also a variety of differences between biobank cohorts and analytic procedures.

We were motivated to examine the effect of controlling for disease status because of its use in the analysis of lab traits in BioBank Japan^10^. Controlling for diseases or risk factors such as tobacco use is a common practice^28^. We considered testing the effect of each disease individually but discarded it as overly cumbersome. Our strategy reflects a broad-spectrum approach in which diagnoses that are rare or have no significant effect on a lab can be rationalized as not causing harm by remaining in the model. The effect of controlling for disease status can be unpredictable. For example, within the components of a lipid panel, controlling for disease status led to a net improvement for HDL catalog SNPs, a net worsening for LDL catalog SNPs, and had cohort-specific impact on triglycerides. From a methodological standpoint, this argues for performing association analyses with and without disease status. From a practical standpoint, the absence of diagnostic data should not be seen as precluding use of a data set.

A drawback of studying clinical labs in real-life cohorts is that some measurements will be artificially modified by prescription medication. We were unable to formally address the effect of medication on genetic analysis because of unreliable measurements of medication. However, it remains an important consideration for future EHR-based lab studies and requires further study. There was indication that in situations where a disease diagnosis is likely to be accompanied by medication, for example a diagnosis of dyslipidemia with lipid labs, controlling for disease status diagnosis serves as a reasonable proxy to treatment status. As research interest in EHR phenotypes increases, we anticipate improved capture of prescription data to facilitate the effects of medications.

A further limitation of this study is the number of analyzed genetic variants. The study was restricted to ∼800K SNPs because BioVU imputed genotypes were unavailable at time of analysis. Although this certainly limited our ability to discover novel variation, the number of SNPs was more than sufficient to perform the primary purpose of the paper, a proof-of-principle replication analysis across a broad range of clinical labs, and the investigation of analytic strategies. However, there are likely many loci remaining to be discovered for these labs, particularly the understudied traits.

In conclusion, we report the first lab-wide genome-wide association study linking data between two independent EHR-based cohorts. We achieved a high degree of replication of prior associations and report a modest number of new associations. In melding these data sets, we addressed key questions in design and analysis of ‘real world’ data that are increasingly relevant.

## Data Availability

Data cannot be shared publicly due to patient confidentiality. The data underlying the results presented in the study are available from University of Michigan Medical School Central Biorepository at https://research.medicine.umich.edu/our-units/central-biorepository/get-access for researchers who meet the criteria for access to confidential data. The meta-analysis summary statistics are available at http://pheweb.sph.umich.edu/mgi-biovu-labs.

http://pheweb.sph.umich.edu/mgi-biovu-labs

## Acknowledgements

The authors acknowledge the University of Michigan Precision Health Initiative and Medical School Central Biorepository for providing biospecimen storage, management, processing and distribution services and the Center for Statistical Genetics in the Department of Biostatistics at the School of Public Health for genotype data curation, imputation, and management in support of this research.

We thank Sebastian Zöllner for valuable feedback on initial drafts.

## Notes

### Competing Interest Statement

G.R.A. is an employee of Regeneron Pharmaceuticals. He owns stock and stock options for Regeneron
Pharmaceuticals. L.A.B and J.C.D receive royalty payments from Nashville Biosciences, a Vanderbilt University Medical Center owned entity.

### Funding Statement

The Michigan Genomics Initiative was supported by institutional funding.

